# Validation of a rapid potency assay for cord blood stem cells using phospho flow cytometry: the IL-3-pSTAT5 assay

**DOI:** 10.1101/2022.03.16.22272489

**Authors:** Carl Simard, Diane Fournier, Patrick Trépanier

## Abstract

Public cord blood banks (CBBs) are required to measure cord blood units (CBUs) potency before their release, allowing for the identification of units that may be unsuitable for hematopoietic transplantation. We have developed a rapid flow cytometry assay based on the measurement of STAT-5 phosphorylation of CD34+ stem cells in response to IL-3 stimulation: the IL-3-pSTAT5 assay.

To adapt the assay from a research setting to its implementation within our CBB regulated operations, we proceeded with a full method validation and a correlation comparison of the IL-3-pSTAT5 assay results with the colony-forming unit assay (CFU) results. A total of 60 CBUs cryopreserved in vials were analyzed by flow cytometry to determine the sensitivity, specificity, intra-assay precision, robustness, reproducibility and inter-laboratory agreement of the assay. The CFU assay was also done on the same samples for comparison purposes.

The assay threshold was established at 50% CD34+CD45+pSTAT5+, which provides a 100% sensitivity and a 98.3% specificity. An average intra-assay CV of 7.3% was determined. All results met our qualitative results acceptance criteria regarding the inter-user and inter-laboratory agreements, IL-3 stimulation time, post-thaw incubation delay and staining time. The IL-3-pSTAT5 assay results correlated well with the total CFU determined using the CFU assay (r^2^ = 0.82, n = 56).

This study shows that our rapid flow cytometry assay can be successfully validated and that the potency data obtained display good sensitivity, specificity and robustness. These results demonstrate the feasibility of implementing this assay within CBB operations, as a validated CB potency assay.

## Introduction

Public cord blood banks (CBBs) are responsible for providing information regarding the characteristics and quality of their cord blood units (CBUs) inventory. Beyond HLA typing, selection of a CBU by a transplant center is usually based on non-functional cellular metrics, such as the total nucleated cell content and CD34+ stem cell count and viability[1,2]. While these criteria may provide reliable information on potential transplant outcome[3], they cannot prevent the selection of units with suboptimal biological activity deceptively displaying satisfactory phenotypic metrics[4]. For that reason, the use of a functional assay targeting the defining characteristics of hematopoietic stem cells, such as the colony-forming unit (CFU) assay, is critical in the selection of CBUs for transplant.

As biological product providers, and with respect to regulatory requirements and inventory and cryopreservation processes performance evaluation, CBBs have to demonstrate that a cryopreserved unit is potent, and must do so using a functional assay. Consequently, most laboratories use the colony-forming unit assay to assess CBU potency. The CFU assay is a reliable indicator of hematopoietic stem cells proliferation and differentiation, and a good predictor of engraftment[5–7]. The principal drawbacks of the CFU assay are its known interlaboratory variability[8,9], the labor intensive nature of the assay and the 7 to 14 days of cellular growth required before getting the results, therefore delaying the release of a unit. This delay can be inconvenient at best, but can also go against the patient’s best interest when an urgent release is required. The development of an alternative assay that is quicker, simpler, and that can be stringently validated intra and interlaboratory remains an unmet need for the stem cells transplant community[10,11].

We previously developed and reported a new flow cytometry method for determining CBU potency within 24 hours, defining the capacity of CD34+CD45+ cells to grow by measuring their level of STAT5-phosphorylation following a short IL-3 stimulation[12]. We demonstrated that the IL-3-pSTAT5 method provided advantages over other currently available alternatives, such as speed, simplicity and good sensitivity. Following the recommended best practices for new potency assay development, we proceeded with a validation of the IL-3-pSTAT5 method in order to implement it within our public CBB operations, as a routine potency test. We herein present the results of the validation regarding the assay sensitivity and specificity, optimal threshold determination, robustness, reproducibility and inter-laboratory agreement. We also present the correlation between the CFU predictive capacity of the IL-3-pSTAT5 method and the total actual CFU measured using the standard CFU assay.

## Materials and methods

### Sample preparation

This study has been approved by Héma-Québec’s Research Ethics Committee and all participants signed an informed consent. Units used in this study were not eligible for banking due to limited volume (< 85 mL) or cell dose (< 1.5 x10^9^ total nucleated cells). CBUs were volume-reduced and cryopreserved following Héma-Québec’s Standard Operating Procedures (SOPs). Plasma and red blood cell extraction was done using an automated blood component separator (Macopresse, MacoPharma, France), reducing CBU volume to between 22 and 26 mL. The cryopreservation solution contained dextran 40 (4.85%) and DMSO (56%) (CryoSure, WAK-Chemie Medical GmbH, Steinbach, Germany) and was used at a final concentration of 10% DMSO. Samples were aliquoted in 1ml cryotubes before being frozen in a CryoMed™ controlled-rate freezer (model 7450, Thermo Scientific, Marietta, OH, USA) at a rate of 1°C/min, and then stored into a vapor-phase liquid nitrogen tank. A standard CBU sample that was processed and cryopreserved according to SOPs and not exposed to a voluntary warming event will be referred to as ‘‘normal’’, while the same sample processed in the same way but exposed to an induced severe warming event post-cryopreservation (from cryopreserved state to room temperature for 24 hours), will be referred to as ‘‘abnormal’’ in this study. The normal samples are expected to score equal or more than the IL-3-pSTAT5 positivity threshold, while the abnormal samples are expected to score under the threshold.

### Flow cytometry

Samples were thawed using an automated thawing system (Thawstar, Biolife Solutions, Bothell, WA, USA). Negative controls were prepared by keeping thawed samples at room temperature for 24h before staining and analysis. Samples were analyzed as previously described[12]. Briefly, 25 µl of thawed cord blood was diluted with 85 µl of Iscove modified Dulbecco’s medium (IMDM) supplemented with 10% fetal bovine serum (FBS; GIBCO, Grand Island, NY, USA) in a 96-well plate and cells were allowed to recover for 45 minutes at 37°C, 5% CO_2_. For each sample, two wells were used as unstimulated controls and two wells were stimulated by adding 1 µl of 10 µg/ml recombinant human interleukin-3 (IL-3; Stem Cells Technologies, Vancouver, Canada). Plates were incubated 20 minutes at 37°C, 5% CO_2_. Samples were then transferred in tubes containing 1.9 ml of prewarmed Phosflow Lyse/Fix buffer (BD Bioscience, Mississauga, Canada) and incubated for 10 minutes in a 37°C water bath. Following centrifugation at 500g for 8 minutes, 1.9 ml of supernatant was removed and 500 µl of 90% methanol-10% PBS was added, before permeabilizing at room temperature for 20 minutes. After centrifugation at 500g for 8 minutes, cell pellets were washed with PBS – 1% BSA and centrifuged again. Cell pellets were suspended in 50 µl of PBS – 1% BSA, mixed with 10 µl of CD45-FITC/CD34-PE mix (BD Biosciences, Mississauga, Canada) and 10 µl of anti-STAT5 (pY694) Alexa Fluor 647 (BD Biosciences, Mississauga, Canada). Staining was done overnight at 4°C. Tubes were centrifuged at 500g for 8 minutes and cell pellets suspended in PBS – 1% BSA. Flow cytometry analyses were performed on Accuri™ C6 flow cytometer (BD, Franklin Lakes, NJ, USA) and analyzed using BD Accuri™ C6 software. For each sample, the Stat5(pY694) positive gate was set to obtain a mean between 2% and 5% of Stat5(pY694) positive CD34 cells for the unstimulated samples. The level of positivity was then determined as the mean of the percentages (rounded to the nearest percent) of IL-3-responsive CD34 cells in the two stimulated samples. A flow chart of the protocol and gating is available in Figure 1.

**Figure 1.**
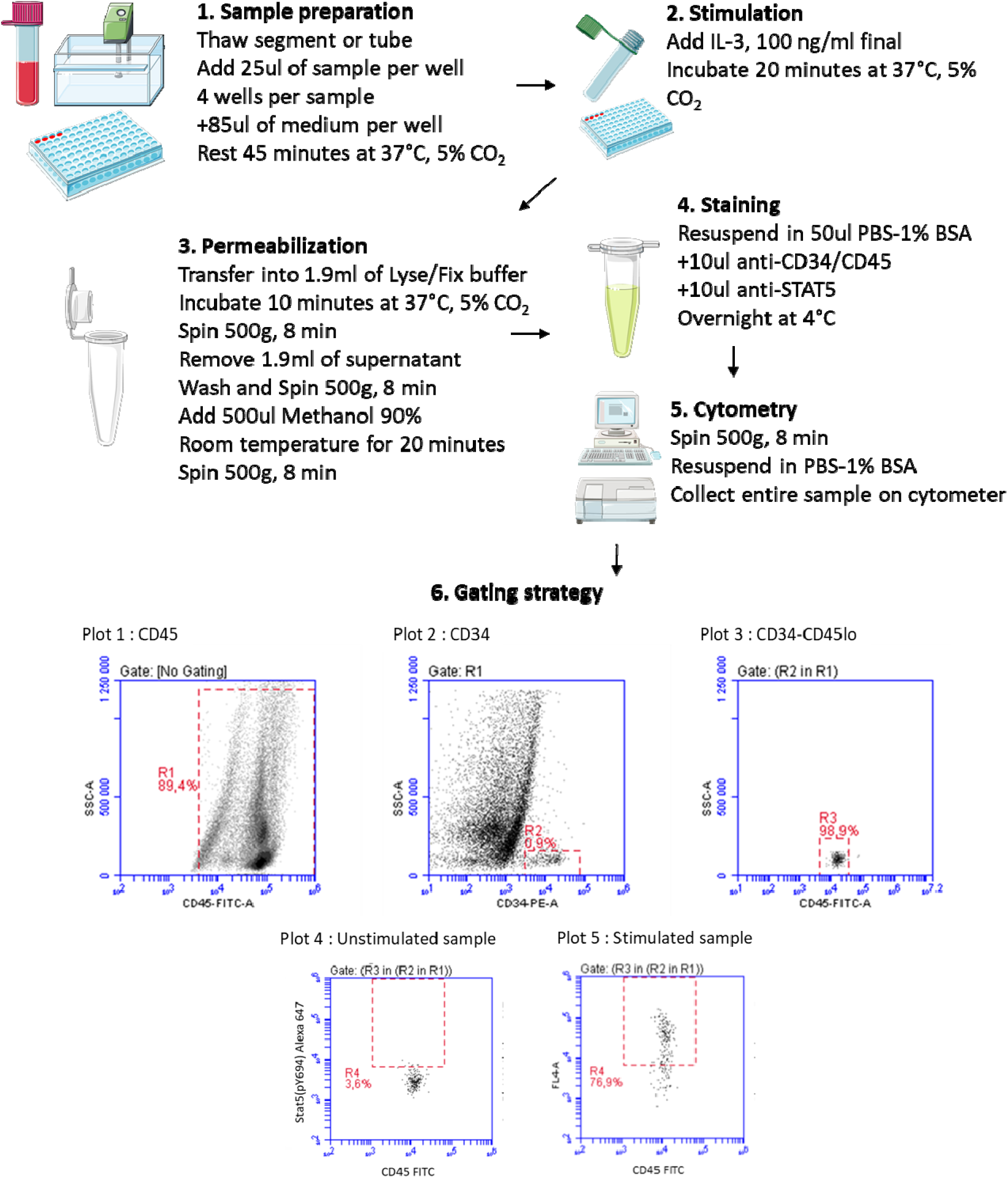
Flowchart of the IL-3-pSTAT5 assay. The assay was divided in five main technical steps, including data collection on the flow cytometer. Step 6 describes the gating strategy. Art used in steps 1 to 5 are from Servier Medical Art (smart.servier.com).

### Colony-forming unit assay

The CFU method was performed according to the manufacturer’s instructions (Stemcell Technologies, British Columbia, Canada), with the modification that thawed samples were diluted with four volumes of Plasmalyte A–5% Human Serum Albumin (HSA). Using CD34 counts before freezing, 1100 CD34 cells were diluted in Iscove modified Dubecco’s medium (IMDM) supplemented with 1% fetal bovine serum in a total volume of 400 μL. From this dilution, 300 μL of cell suspension was mixed with 3 mL of methylcellulose-based medium (Methocult Optimum H4034, Stemcell Technologies) before plating 1.1 mL (250 CD34 cells) in duplicate 30-mm Petri dishes. CFUs were counted manually after 14 days of incubation at 37°C, 5% CO_2_.

### Data analysis

Receiver Operating Characteristic (ROC) curve analysis, specificity, sensitivity and threshold values were determined using package OptimalCutpoints 1.1-4[13] in R 4.0.3 and Graphpad Prism 9 (GraphPad Software, San Diego, CA, USA). Confidence intervals were calculated in R using the epiR 2.0.19 package and the Bland-Altman analysis with blandr 0.5.1.

## Results

### Flowchart of the IL-3-pSTAT5 assay

The assay steps were optimized, simplified, and are presented in Figure 1 as a flowchart. The plots 1 to 3 in part 6 of the Figure 1 show the gating strategy to isolate the CD34+ stem cells. The plot 4 shows the gate placement using an unstimulated sample, and the plot 5 shows the positive results obtained with IL-3 stimulated cells.

### Assay specificity and sensitivity

Specificity and sensitivity were assessed by analyzing normal (n=60) and abnormal samples (n=60), for a total of 120 samples. The results’ graphical representation, the ROC curve and the sensitivity vs specificity curve are presented in Figure 2A, B and C, respectively. The graphical representation in Figure 2A shows the distribution of normal and abnormal samples relative to the 50% threshold. The ROC curve in Figure 2B shows the optimal threshold point of 48,7%, based on Youden’s index. Sensitivity and specificity curves were plotted against assay threshold values in Figure 2C. The assay threshold was rounded and established at 50%, which provides a sensitivity of 100% (CI95 94.0% - 100%) and specificity is 98.3% (CI95 91.2% - 100%). This 50% positivity threshold was used for the entire method validation: an IL-3-pSTAT5 value equal to or greater than 50% was interpreted as ‘‘pass’’, and any value under 50% as ‘‘fail’’.

**Figure 2:**
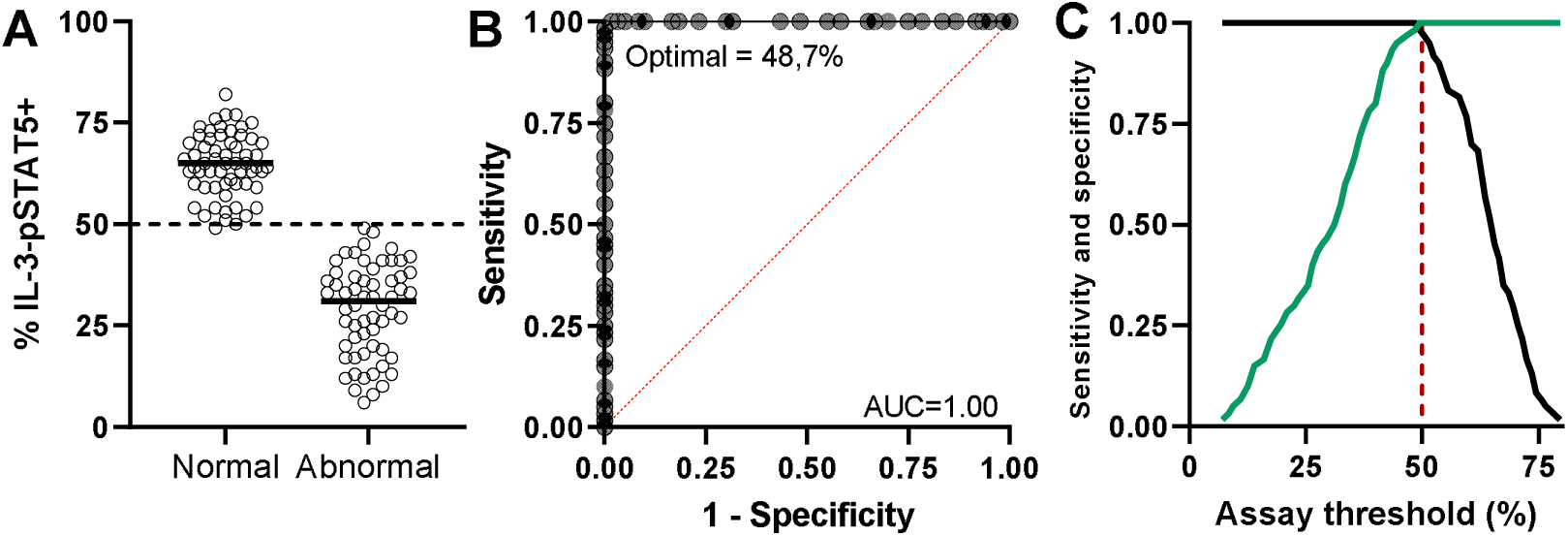
A) Distribution of results for normal (n = 60) and abnormal (n = 60) samples. The dotted line corresponds to the established positivity threshold of 50% IL-3-responsive CD34+ cells. The black line within the dots is the median. B) ROC curve with optimal threshold point based on Youden’s index and the line of no discrimination (dotted, red). C) Sensitivity (green) and specificity (black) curves relative to the assay threshold values. The red dotted line corresponds to the threshold value of 50% used for the validation. At this threshold, sensitivity is 100% (CI95 94.0% - 100%) and specificity is 98.3% (CI95 91.2% - 100%).

### Intra-assay precision

Samples from three different CBUs were tested ten consecutive times by the same operator, using the same cytometer in order to measure the assay’s coefficient of variation (CV). As shown in table 1, sample series had CVs of 7.2%, 9.8% and 4.8%, with a mean of 7.3%. All analyses met our ≤10% CV acceptance criteria.

**Table 1:** Intra-assay precision statistics for three independent samples.

| Sample | N | Mean %IL-3 | SD | CV | Min | Max |
| --- | --- | --- | --- | --- | --- | --- |
| **1** | 10 | 64.9 | 4.7 | 0.072 | 57.9 | 68.2 |
| **2** | 10 | 59.6 | 5.9 | 0.098 | 52.1 | 72.9 |
| **3** | 10 | 46.9 | 2.3 | 0.048 | 42.5 | 49.7 |
| **Mean** | 30 | 57.1 | 4.3 | 0.073 | 50.8 | 63.6 |

### Reproducibility and robustness

The robustness of the assay was evaluated by analyzing normal and abnormal samples from ten CBUs with varying parameters; different operators (Figure 3A), IL-3 stimulation time (15 and 25 minutes; Figure 3B), post-thaw incubation time (30 and 60 minutes; Figure 3C), staining time (24 and 72 hours; Figure 3D), and different laboratories (Figure 3E). The inter-user results (Figure 3A) and varying IL-3 stimulation time results (Figure 3B) show a correct qualitative match of 7 pass and 3 fail samples for both sets of parameters. Post-thaw incubation time results (Figure 3C) show a correct qualitative match of 4 pass and 6 fail samples for both 30- and 60-minutes incubation periods. The results from staining time of 24h vs 72h (Figure 3D) show qualitative results of 6 pass and 3 fail samples, with one sample (#6) ‘‘pass’’ at 24h and ‘‘fail’’ at 72h. Finally, the inter-laboratory agreement (Figure 3E) was done by assaying ten CB samples in two different laboratories. Data show that both laboratories identified the 7 normal and 3 abnormal samples correctly. All results met our <10% discordant qualitative results acceptance criteria.

**Figure 3:**
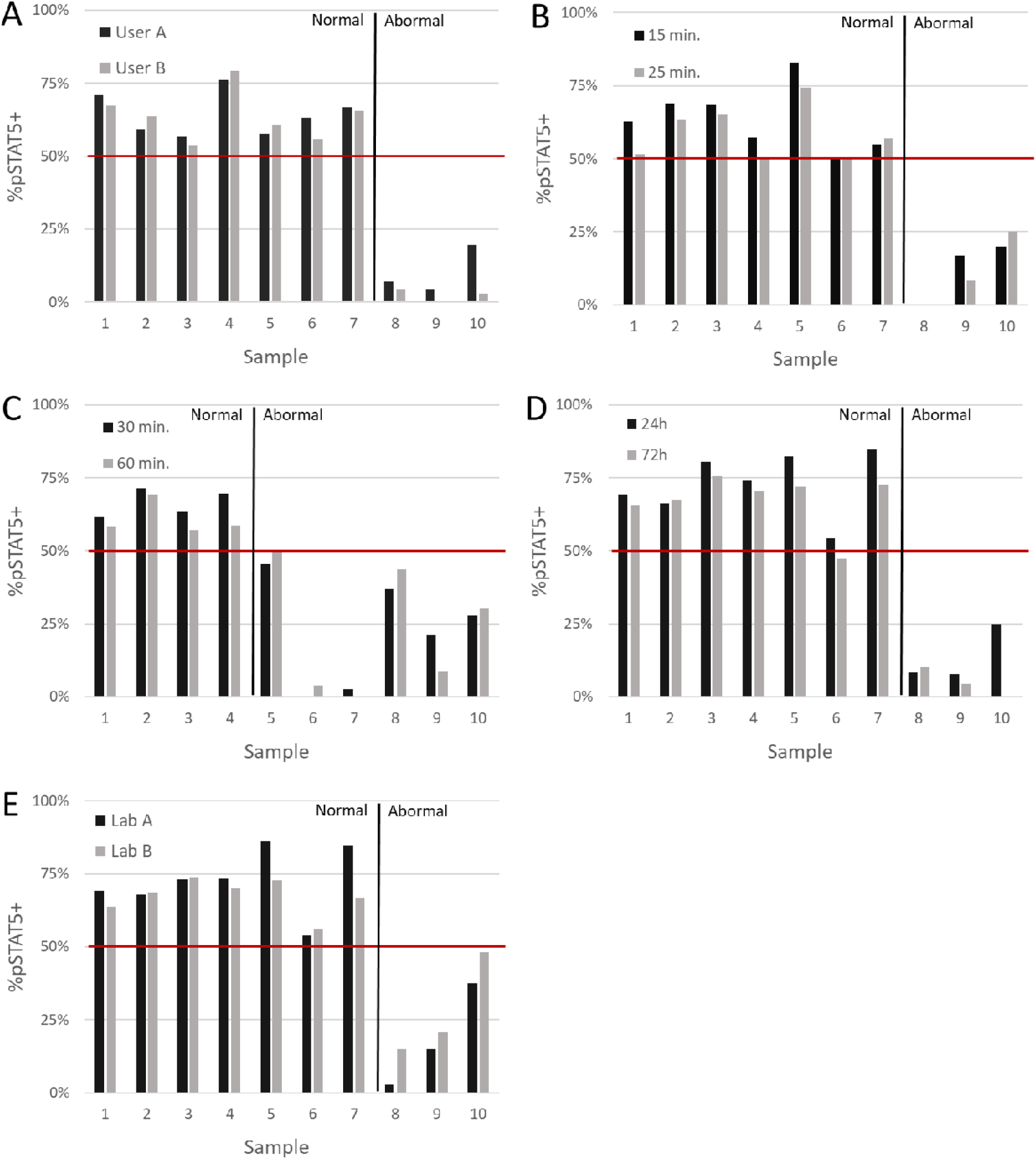
Robustness of the IL-3-pSTAT5 technique to variation in A) users, B) IL-3 stimulation time (15 vs 25 minutes), C) resting time after thawing (30 vs 60 minutes), D) staining time (24 vs 72 hours) and E) inter-laboratory agreement. The red horizontal line represents the 50% positivity threshold. The black vertical line separate normal samples (expected conformant, ≥50% IL-3-pSTAT5 result) from abnormal ones (expected non-conformant, <50% IL-3-pSTAT5 result). Sample numbers (1 to 10) were randomly chosen and do not correspond to the same samples in figures A to E. Samples were transported between laboratories using a validated dry shipper and distribution protocol.

### Correlation with total CFU

In addition to the IL-3-pSTAT5 assay, 14-day CFU assays were performed on the same samples (N=56). Total CFU count results from the CFU assay were plotted against a CFU-estimate calculated from IL-3 potency results. This CFU-estimate is calculated by multiplying the percentage of CD34+CD45+pSTAT5+ cells with the number of postthaw CD34 cells (%IL-3*post-thaw CD34 count). The correlation between both total CFU and CFU-estimate is good with an r^2^ of 0.82 (Figure 4A). The Bland-Altman plot (Figure 4B) shows an average bias towards overestimation of 246 164 CFU/units, meaning that samples with less than 250 000 estimated CFU are expected to show very little to no growth in the CFU assay.

**Figure 4.**
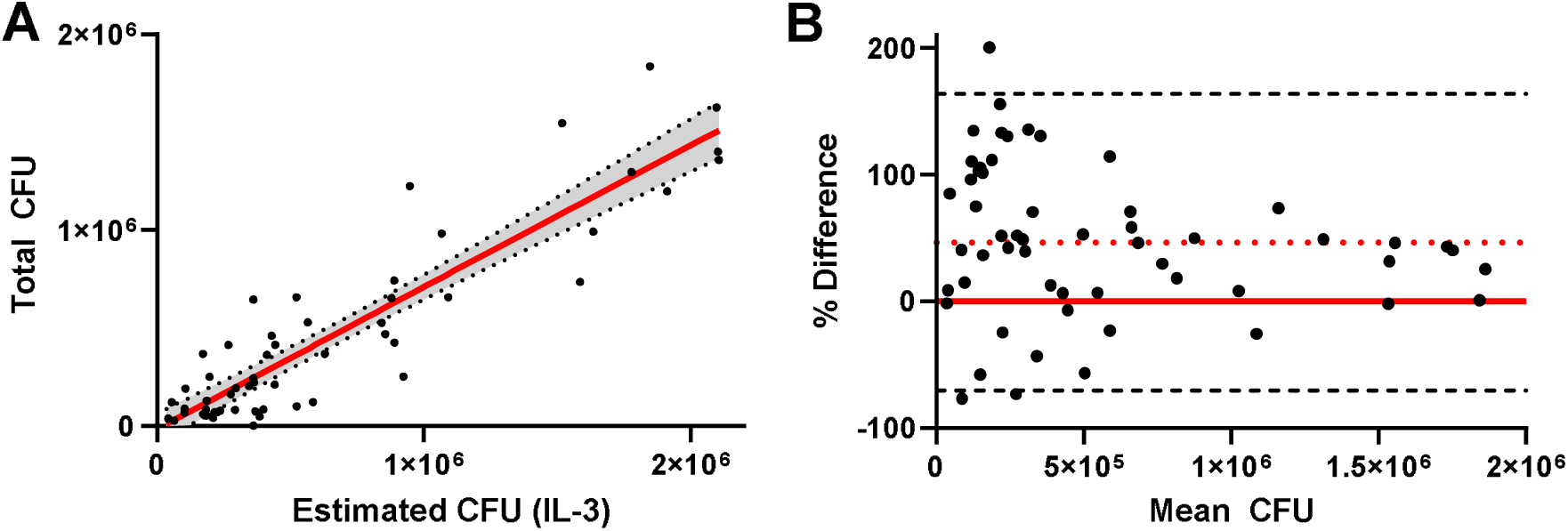
Comparison of the estimated CFU number derived from IL-3-pSTAT5 assay results and total CFU from CFU assay. A) Linear regression (red line) and 95% confidence interval (shadowed region between dotted lines) between the number of CFU estimated from the IL-3-pSTAT5 assay and that measured by CFU assay culture (n = 56, r^2^ = 0.82). B) Bland-Altman plot (n = 56) of the relative differences between the estimated CFU numbers and actual CFU numbers, the red line representing no difference, the dotted red line being the mean bias (46.4%) and the dashed lines corresponding to the limits of agreement (165.8% and -73.1%). Samples with more than 250 000 events have a bias of 31.1% and limits of agreement of 125.9% and – 63.7%. All data are for total CFU content of a CBU.

## Discussion

This work presents the full validation of a novel, rapid and optimized potency assay for CB stem cells using intracellular flow cytometry to measure STAT5 phosphorylation in CD34+CD45+ cells following IL-3 stimulation. The assay sensitivity and specificity at the optimal threshold and the assay’s robustness, reproducibility and inter-laboratory agreement were determined and proven satisfactory based on pre-established validation requirements. Beyond the IL-3-pSTAT5 assay’s qualitative capacity to discriminate between a normal (conformant) and an abnormal (non-conformant) CBU, results were shown to correlate with total CFU count. This suggests the assay could be used not only as a potency and release test, but also as a predictor of CFU results, in cases where the information is required urgently. In situations where colony differentiation counts or exact CFU values are needed, the standard CFU assay should still be performed.

Our IL-3-pSTAT5 assay meets the many expectations for next-generation potency testing, such as automated testing, high throughput and rapid turnover time. The marginal cost of introducing this technique in a laboratory that already performs flow cytometry analysis on cord blood is low, and the reagent cost is comparable to that of the CFU assay. Nevertheless, the IL-3-pSTAT5 assay in its current and validated form does not provide information regarding the differentiation state of stem cells, which can be observed with the classical CFU assay, albeit after many days of culture. Although this assay may appear less informative than the CFU assay, this was done by design, as using longer and more complex cytokine stimulations, cultivating cells over many days and adding different antibodies defeated the purpose of a quick, simple and easy to implement potency assay to use in our CBB operations. The IL-3-pSTAT5 validated qualitative assay was shown to be the quickest way for our CBB to reliably and robustly evaluate CBU potency, all while resolving the challenges of the CFU assay’s variability and lower sensitivity, as we previously described[12]. In addition, it is known that proficiency testing results are variable between laboratories, with efforts conducted by the National Marrow Donor Program (NMDP) to address the issue. On this point, our approach should circumvent the presence of contaminating RBC, and get around other differences between processing methods which can lead to artefacts interfering with optimal flow cytometry analysis and conclusions[14].

While our validation does not guarantee the assay will easily be made functional in any other stem cells laboratories willing to use it, an international multicenter study is ongoing and should address any remaining concern. We hope that such use of intracellular flow cytometry will help circumvent current challenges associated with potency testing, and allow for faster, more reliable release of products to clinical programs, consequently serving the patient’s best interest.

## Data Availability

All data produced in the present work are contained in the manuscript

## Abbreviations

(CFU): Colony-forming unit
(CB): Cord Blood
(CBB): Cord Blood Bank
(CBU): Cord Blood Unit
(FACT): Foundation for the Accreditation of Cellular Therapy
(PBSC): Peripheral Blood Stem Cells
(SOP): Standard Operating Procedure
(WMDA): World Marrow Donor Association

## Author Contributions

Conception and design of the study: PT, DF and CS. Acquisition of data: CS. Analysis and interpretation of data: PT and CS. Drafting and revising the manuscript: CS, PT and DF. All authors have approved the final article.

## Highlights

- Current cord blood stem cells potency assays are challenging and results variable
- The novel IL-3-pSTAT5 assay can rapidly and reliably determine stem cells potency
- The method was fully and successfully validated, meeting all validation criteria
- The IL-3-pSTAT5 correlates well with the colony-forming unit assay
- This quick and validated assay could be an important tool for cord blood banks

## Acknowledgements

The authors thank all the mothers who have contributed to both the establishment of the Héma-Québec Public Cord Blood Bank and to this study with their generous donations, and are grateful to Héma-Québec’s dedicated Cord Blood Bank team. The authors would like to thank Dr Sonia Néron for her important contribution to this work and Marie-Ève Rhéaume for revising the manuscript.

